# Clinical Performance of the Alinity m HR HPV Assay in a Large Urban Cervical Cancer Screening Program in the United States

**DOI:** 10.1101/2024.02.12.24302707

**Authors:** Joshua Kostera, Almedina Tursunovic, Paige Botts, Yan Zhang, Regina Galloway, April Davis, Tong Yang

## Abstract

**Background:** Certain high-risk human papillomavirus (HR HPV) genotypes may carry greater risk of progression to high-grade cervical cancer, and extended HR HPV genotyping may help refine risk assessment.

**Objectives:** To examine the performance of the Alinity m HR HPV assay in comparison to cobas 4800 HPV assay (cobas 4800) and the association between cytology results, age, and extended HR HPV genotyping.

**Study design:** This was a cross-sectional cohort study of 149 de-identified remnant cervical cytology specimens collected in ThinPrep as part of a routine cervical cancer screening program at a large urban hospital system. Specimens were tested on the Alinity m HR HPV assay and the cobas 4800 assay; results were analyzed by age and cytology.

**Results:** Overall percent agreement (OPA) between Alinity m and the cobas 4800 assay was >96% for the detection of any HR HPV, HPV 16 and 18, and other HR HPV genotypes, across all cytology categories (Cohen’s κ ≥0.91). The majority of Alinity m negative results had negative for intraepithelial lesions or malignancy (NILM) or atypical squamous cells of undetermined significance (ASC-US) cytology. Alinity m positivity, particularly for HPV 16, 18, and Other A (HPV 31, 33, 52, and 58), increased with the severity of atypical cytology. Alinity m positivity was more often associated with ASC-US cytology in specimens from individuals ≤29 years of age and with low-grade squamous intraepithelial lesion (LSIL) cytology or more severe cervical disease in individuals ≥30 years of age.

**Conclusions:** The extended genotyping capability of the Alinity m HR HPV assay may aid in the detection of specimens with increased risk of progressing to high-grade cervical disease.

## Introduction

High-risk human papillomavirus (HR HPV) testing is recommended by the American Cancer Society (ACS) as well as the American Society for Colposcopy and Cervical Pathology (ASCCP) [1]. HPV testing has become a significant component of cervical cancer screening programs in recent years, mirroring a reduction in cervical cancer mortality rate in the United States [2]. Persistent infection with one or more HR HPV genotypes (HPV 16, 18, 31, 33, 35, 39, 45, 51, 52, 56, 58, 59, 66, and 68) is recognized as one of the major causes of viral-related cancers in both men and women, and its strong association with cervical cancer is well established [3,4]. The majority of HPV infections are sub-clinical, transient, and resolve spontaneously within 2 years; however, approximately 10% of HR HPV infections persist, with an elevated risk for progression to high-grade cervical intraepithelial neoplasia (CIN) and the development of cervical cancer [5, 6].

Outcomes from recent large-scale clinical trials, meta-analyses, and real-world clinical evaluations have found that HPV genotypes beyond 16 and 18 carry a risk for high-grade CIN (≥CIN3) that meet or exceed the current risk threshold for referral to colposcopy. Specifically, these studies have shown that HPV 31, 33, 35, 45, 52, and 58 are closely associated with an intermediate-to-high ≥CIN3 risk, and HPV 31 and 33 are associated with an accelerated rate of disease progression, similar to the association between HPV 18 and 45 infection and high-grade cervical disease [7-10]. Additionally, studies suggest that the lowest risk HR HPV genotypes (39, 51, 56, 59, 66, and 68) carry a relatively lower ≥CIN3 relative risk, and patients may be counseled to retest after a set period of time rather than be referred to colposcopy [10-17]. The use of extended HR HPV genotyping to stratify cervical cancer risk supports the most recent ASCCP guideline update [18], which advocates for a risk-based approach to identify high-grade CIN during screening in order to prevent cervical cancer [18,27].

Here, we compared the clinical performance of the recently FDA approved Alinity m HR HPV assay (Abbott Laboratories, Abbott Park, IL) to the cobas 4800 assay (Roche, Basel, Switzerland). The extended HR HPV genotype analysis included comparative sensitivity and specificity along with age-related prevalence of HPV infection by Alinity m HR HPV results and cytology in our study cohort.

## Methods

### Setting and Participants

Remnant non-sequential de-identified patient specimens, collected as part of the Ochsner Health routine cervical cancer screening program, were selected by the Ochsner Health molecular pathology laboratory for this study. Specimens were chosen to include a wide range of cytological classifications with HPV positive and negative results. Specimens were collected in ThinPrep® LBC medium and processed for cytology. Specimens were aliquoted for molecular testing on the Alinity m HR HPV assay. HPV results that were discordant between Alinity and cobas 4800 were tested with the Aptima HPV detection assay first and the positive samples were then reflexed to the Aptima 16, 18/45 GT assay. Patient identifiers were removed from the specimens and the study was conducted in accordance with an approved Ochsner Health Institutional Review Board (IRB) protocol.

### Cytology

Cytology was performed using a ThinPrep® 2000 processor and specimens were graded based on the 2014 Bethesda System. Cytology results were classified as negative for intraepithelial lesions or malignancy (NILM), atypical squamous cells of undetermined significance (ASC-US), or as having any atypical cytology beyond ASC-US (≥LSIL), including low-grade squamous intraepithelial lesion (LSIL), atypical glandular cells (AGC), high-grade squamous intraepithelial lesion (HSIL), Atypical squamous cells, cannot exclude HSIL (ASC-H), and squamous cell carcinoma or adenocarcinoma. All results were reviewed by a cytotechnologist and all abnormal cases and selected negative cases (including random 10% and negative cases with previous abnormal results) were reviewed the pathologist.

### Assays

The Alinity m HR HPV assay (Alinity m) is a qualitative real-time PCR-based assay that simultaneously amplifies and detects genotypes HPV 16, HPV 18, and HPV 45 while reporting the 11 other HR HPV genotypes in 2 aggregates: Other HR HPV A (31/33/52/58) and Other HR HPV B (35/39/51/56/59/66/68). The assay is approved with ThinPrep and SurePath specimen types, includes a cellular control (CC) to ensure specimen adequacy and sample extraction and amplification efficiency.

The qualitative cobas HPV assay is FDA-approved for primary screening and co-testing with cytology, detects all 14 HR HPV genotypes and differentiates between HPV 16, 18, and the remaining 12 other genotypes (HR HPV Other) [19]. The test is approved with ThinPrep and SurePath specimen types, includes a cellular control and is run on the automated cobas 4800 system.

The Aptima HPV qualitative assay is only FDA-approved for co-testing with cytology that detects all 14 HR HPV genotypes, but does not differentiate between them, reporting only a positive or negative assay result [20]. Specimens positive on the Aptima HPV assay were run on the Aptima HPV 16 18/45 GT assay [21]. Both assays are only approved with the ThinPrep specimens which are run on the Panther system.

### Statistical analysis

Overall, negative, and positive percent agreement rates (OPA, NPA, and PPA) were determined for the Alinity m and cobas assays, for specimens determined to have NILM, ASC-US, or ≥LSIL cytology, for all specimens positive for HR HPV and separately for the clinically relevant HPV 16 and 18 genotypes and the other non-HPV 16/18 genotypes. Cohen’s kappa analysis was performed to compare overall agreement between the tests for each cytology category. Age-related data associated with an Alinity m result across cytology categories NILM, ASC-US, and ≥LSIL were also examined in this cohort. Data analyses were performed using SAS software version 9.3 or higher (SAS, Cary, NC).

## Results

### Participants

A total of 149 de-identified residual cervical clinical specimens collected in ThinPrep® were included in the study. Fifty-six specimens were NILM and 93 samples were ≥ASC-US by cytology. The distribution of cytology and hierarchical Alinity m and cobas 4800 assay results are shown in Tables 1 and 2. Most specimens in this cohort with atypical cytology were ASC-US (60%, n=56/93) or LSIL (30%, n=28/93). All specimens tested in this cohort generated valid results with the Alinity m and cobas 4800 assays.

**Table 1.** Cytology Distribution by Hierarchical HR HPV Genotype on Alinity m HR HPV Assay.

| Cytology Result | HPV16 | HPV18 | HPV45 | Other HR HPV A | Other HR HPV B | HR HPV Detected | Not Detected | Total |
| --- | --- | --- | --- | --- | --- | --- | --- | --- |
| Adenocarcinoma | 0 | 0 | 0 | 0 | 0 | 0 | 1 | 1 |
| AGC | 0 | 0 | 0 | 0 | 2 | 2 | 2 | 4 |
| HSIL | 1 | 0 | 3 | 0 | 0 | 4 | 0 | 4 |
| LSIL | 2 | 2 | 3 | 8 | 10 | 25 | 3 | 28 |
| ASC-US | 2 | 1 | 0 | 11 | 23 | 37 | 19 | 56 |
| NILM | 1 | 1 | 2 | 8 | 13 | 25 | 31 | 56 |
| Total | 6 | 4 | 8 | 27 | 48 | 93 | 56 | 149 |

**Table 2.** Cytology Distribution by Hierarchical HR HPV Genotype on cobas 4800 HPV Assay.

| Cytology Result | HPV16 | HPV18 | Other HR HPV | HR HPV Detected | Not Detected | Total |
| --- | --- | --- | --- | --- | --- | --- |
| Adenocarcinoma | 0 | 0 | 0 | 0 | 1 | 1 |
| AGC | 0 | 0 | 2 | 2 | 2 | 4 |
| HSIL | 1 | 0 | 3 | 4 | 0 | 4 |
| LSIL | 3 | 2 | 21 | 26 | 2 | 28 |
| ASCUS | 2 | 1 | 37 | 40 | 16 | 56 |
| NILM | 1 | 1 | 21 | 23 | 33 | 56 |
| Total | 7 | 4 | 84 | 95 | 54 | 149 |

### Agreement between Alinity m HR HPV and cobas 4800 assay results

Table 3 shows the agreement between Alinity m and cobas 4800 assay results for HR HPV Detected in ≥LSIL, ASC-US, NILM, and all categories combined by cytology. The Alinity m results demonstrated very good agreement with the cobas 4800 results across all individual cytological categories with an OPA ≥94.6% and a total OPA of 96.0%.

**Table 3.** Percent Agreement with 95%CI Between Alinity m HR HPV and cobas 4800 HPV Assay Results for HR HPV Detected, by Cytology.

| Cytology Result | PPA (95% CI) | NPA (95% CI) | OPA (95% CI) | Alinity m Result | cobas Result |  |  |
| --- | --- | --- | --- | --- | --- | --- | --- |
|  |  |  |  |  | HR HPV Detected | Not Detected | Total |
| ≥LSIL | 96.9<br>(84.3,99.4) | 100.0<br>(56.6,100.0) | 97.3<br>(86.2,99.5) | HR HPV Detected | 31 | 0 | 31 |
|  |  |  |  | Not Detected | 1 | 5 | 6 |
|  |  |  |  | Total | 32 | 5 | 37 |
| ASC-US | 92.5<br>(80.1,97.4) | 100.0<br>(80.6,100.0) | 94.6<br>(85.4,98.2) | HR HPV Detected | 37 | 0 | 37 |
|  |  |  |  | Not Detected | 3 | 16 | 19 |
|  |  |  |  | Total | 40 | 16 | 56 |
| NILM | 100.0<br>(85.7,100.0) | 93.9<br>(80.4,98.3) | 96.4<br>(87.9,99.0) | HR HPV Detected | 23 | 2 | 25 |
|  |  |  |  | Not Detected | 0 | 31 | 31 |
|  |  |  |  | Total | 23 | 33 | 56 |
| Total | 95.8<br>(89.7,98.4) | 96.3<br>(87.5,99.0) | 96.0<br>(91.5,98.1) | HR HPV Detected | 91 | 2 | 93 |
|  |  |  |  | Not Detected | 4 | 52 | 56 |
|  |  |  |  | Total | 95 | 54 | 149 |

Percent agreement between Alinity m and cobas assay results for detecting HPV 16 or 18 in specimens with ≥LSIL, ASC-US, NILM, and all categories combined by cytology is shown in Table 4. OPA was calculated to be ≥97.3% across all individual cytological categories with a total OPA at 99.3% for Alinity m vs cobas results.

**Table 4.** Percent Agreement with 95% CI Between Alinity m HR HPV and cobas 4800 HPV Assay Results for HPV 16 or 18 Detected Specimens, by Cytology.

| Cytology Result | PPA (95% CI) | NPA (95% CI) | OPA (95% CI) | Alinity m Result HPV16/18 | cobas Result HPV16/18 |  |  |
| --- | --- | --- | --- | --- | --- | --- | --- |
|  |  |  |  |  | Positive | Negative | Total |
| ≥LSIL | 83.3<br>(43.6,97.0) | 100.0<br>(89.0,100.0) | 97.3<br>(86.2,99.5) | Positive | 5 | 0 | 5 |
|  |  |  |  | Negative | 1* | 31 | 32 |
|  |  |  |  | Total | 6 | 31 | 37 |
| ASC-US | 100.0<br>(43.9,100.0) | 100.0<br>(93.2,100.0) | 100.0<br>(93.6,100.0) | Positive | 3 | 0 | 3 |
|  |  |  |  | Negative | 0 | 53 | 53 |
|  |  |  |  | Total | 3 | 53 | 56 |
| NILM | 100.0<br>(34.2,100.0) | 100.0<br>(93.4,100.0) | 100.0<br>(93.6,100.0) | Positive | 2 | 0 | 2 |
|  |  |  |  | Negative | 0 | 54 | 54 |
|  |  |  |  | Total | 2 | 54 | 56 |
| Total | 90.9<br>(62.3,98.4) | 100.0<br>(97.3,100.0) | 99.3<br>(96.3,99.9) | Positive | 10 | 0 | 10 |
|  |  |  |  | Negative | 1 | 138 | 139 |
|  |  |  |  | Total | 11 | 138 | 149 |

\*cobas 4800 Result, HPV 16 CN = 39.7.

The percent agreement between Alinity m HPV 45, Other HR HPV A, and Other HR HPV B results and cobas HR HPV Other results for extended genotyping in specimens with ≥LSIL, ASC-US, NILM, and all categories combined by cytology is shown in Table 5. OPA was ≥94.6% across all cytological categories with a total OPA at 96.6% for Alinity m vs cobas 4800 results for detection of non-HPV 16/18 genotypes.

**Table 5.** Percent Agreement with 95% CI Between Alinity m HR HPV Assay Results (HPV 45, Other HR HPV A, Other HR HPV B) and cobas 4800 Assay Results (Other HR HPV), by Cytology.

| Cytology Result | PPA (95% CI) | NPA (95% CI) | OPA (95% CI) | Alinity m Result<br>Other HR HPV | cobas Result<br>Other HR HPV |  |  |
| --- | --- | --- | --- | --- | --- | --- | --- |
|  |  |  |  |  | Positive | Negative | Total |
| ≥LSIL | 100.0<br>(87.5,100.0) | 100.0<br>(72.2,100.0) | 100.0<br>(90.6,100.0) | Positive | 27 | 0 | 27 |
|  |  |  |  | Negative | 0 | 10 | 10 |
|  |  |  |  | Total | 27 | 10 | 37 |
| ASC-US | 91.9<br>(78.7,97.2) | 100.0<br>(83.2,100.0) | 94.6<br>(85.4,98.2) | Positive | 34 | 0 | 34 |
|  |  |  |  | Negative | 3* | 19 | 22 |
|  |  |  |  | Total | 37 | 19 | 56 |
| NILM | 100.0<br>(84.5,100.0) | 94.3<br>(81.4,98.4) | 96.4<br>(87.9,99.0) | Positive | 21 | 2<sup>#</sup> | 23 |
|  |  |  |  | Negative | 0 | 33 | 33 |
|  |  |  |  | Total | 21 | 35 | 56 |
| Total | 96.5<br>(90.1,98.8) | 96.9<br>(89.3,99.1) | 96.6<br>(92.4,98.6) | Positive | 82 | 2 | 84 |
|  |  |  |  | Negative | 3 | 62 | 65 |
|  |  |  |  | Total | 85 | 64 | 149 |

\*cobas 4800 Results, Other HR HPV CN's = 34.1, 37.3, 38.2.
<sup>#</sup>Alinity m Results, Other HR HPV B CN's = 27.19, 27.79.

We further analyzed the agreement between Alinity m and cobas results in NILM, ASC-US, ≥LSIL, and all cytology categories combined using Cohen’s kappa analysis. Comparison between the 2 assays showed strong to perfect agreement (kappa values 0.88 to 1.00), with almost perfect agreement across all cytology categories with kappa values calculated to be ≥0.91 (Table 6).

**Table 6.** Cohen’s Kappa Analysis of the Agreement Between Alinity m HR HPV and cobas 4800 Results for HR HPV Detected, HPV 16 or 18, and Other HR HPV Genotypes, by Cytology.

|  | Cytology Result | Estimate | 95% CI |
| --- | --- | --- | --- |
| HR HPV Detected | ≥LSIL | 0.89 | (0.69, 1.00) |
|  | ASCUS | 0.88 | (0.74, 1.00) |
|  | NILM | 0.93 | (0.83, 1.00) |
|  | Total | 0.91 | (0.85, 0.98) |
| HPV 16 or 18 Positive | ≥LSIL | 0.89 | (0.69, 1.00) |
|  | ASCUS | 1 | (1.00, 1.00) |
|  | NILM | 1 | (1.00, 1.00) |
|  | Total | 0.95 | (0.85, 1.00) |
| Other HR HPV Positive | ≥LSIL | 1 | (1.00, 1.00) |
|  | ASCUS | 0.88 | (0.76, 1.00) |
|  | NILM | 0.93 | (0.82, 1.00) |
|  | Total | 0.93 | (0.87, 0.99) |

### Age associated data with cytology distribution by Alinity m HR HPV result

The average age of all patients in this cohort was 40.3 years. The average age associated with an Alinity m result for HPV 16 or 18, HPV 45, Other HR HPV A, and Other HR HPV B across cytology categories is shown in Table 7. The distribution of cytological category and Alinity m Detected results within the age groups of 21-29 years, 30-44 years, and ≥45 years is shown in Figure 1.

**Table 7.** Average Age in Years Associated with an Alinity m HR HPV Result Across Cytology Categories.

| Alinity m HR HPV Result | Cytology |  |  |  |
| --- | --- | --- | --- | --- |
|  | NILM | ASC-US | ≥LSIL | Total |
| HPV 16 or 18 | 47.5 | 40.7 | 39.2 | 42.5 |
| HPV 45 | 39.0 | N/A* | 44.7 | 41.9 |
| Other HR HPV A (31/33/52/58) | 49.1 | 37.0 | 42.2 | 42.8 |
| Other HR HPV B (35/39/51/56/59/66/68) | 37.3 | 34.4 | 37.2 | 36.3 |

\*No Alinity m HR HPV 45 Results were observed with ASC-US cytology.

**Figure 1.**
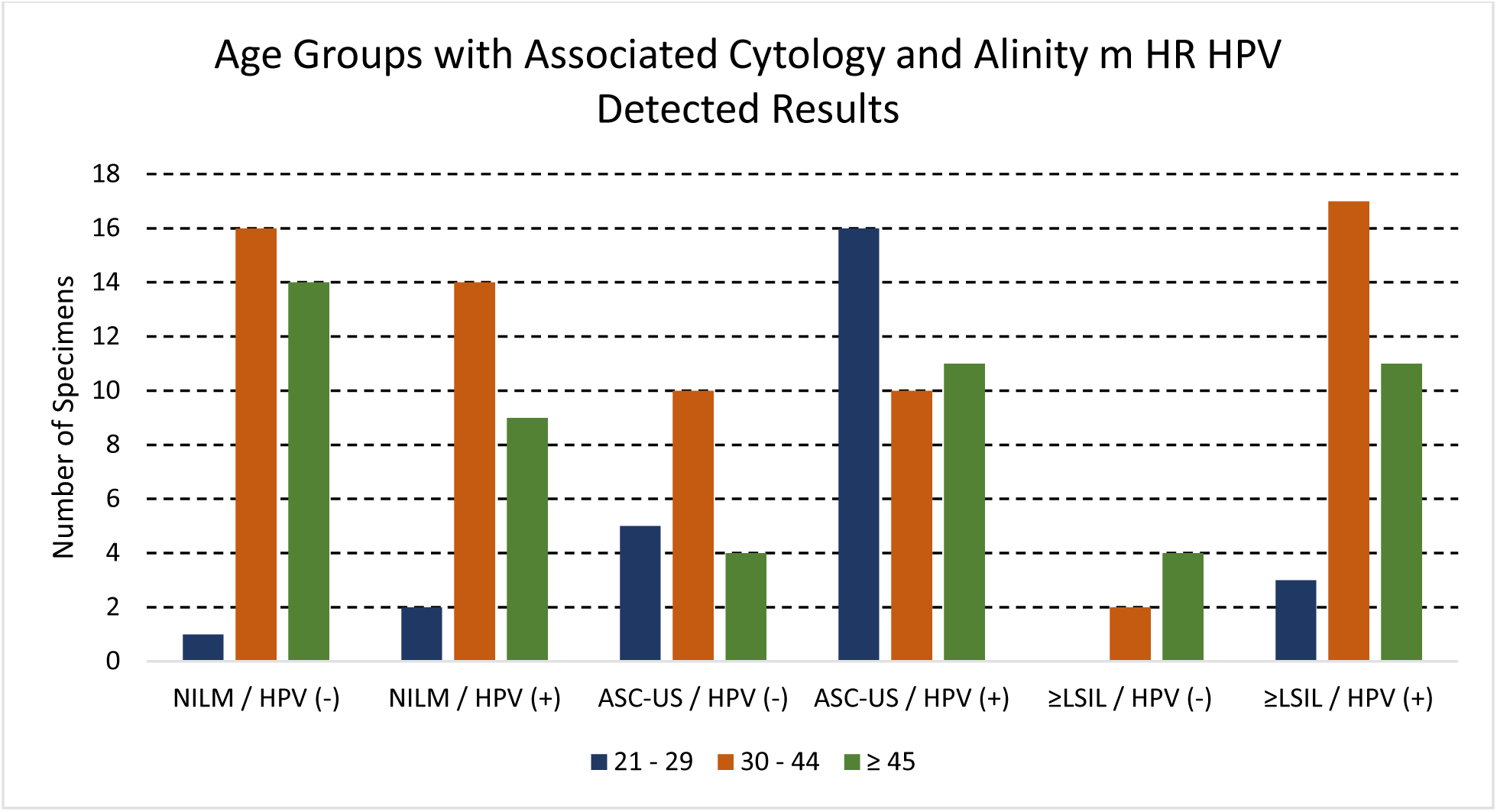
The distribution of cytological category (NILM, ASC-US, ≥LSIL) and Alinity m HR HPV Detected results with associated age groups.

Notably, in the 21- to 29-year age group, approximately 3-fold more specimens with ASC-US cytology had a positive Alinity m result (n=16) versus a negative result (n=5). A comparable pattern was observed for ASC-US specimens in the age group ≥45 years (n=11 Alinity m positive vs. n=4 Alinity m negative). For specimens with ≥LSIL cytology, 8-fold more specimens were Alinity m positive vs. negative in the 30- to 44-year age group (n=17 Alinity m positive vs. n=2 Alinity m negative). In the ≥45-year age group with ≥LSIL cytology, 3-fold more specimens were Alinity m positive than negative (n=11 Alinity m positive vs. n=4 Alinity m negative).

## Discussion

In this study, we found high agreement rates between Alinity m HR HPV and cobas 4800 HPV assay results in specimens with NILM, ASC-US, and ≥LSIL cytology. Cohen’s kappa analysis showed near perfect agreement for all cytology classifications between the 2 assays for clinically relevant HPV 16/18 (κ=0.95), as well as extended genotyping of other HR HPV genotypes (κ=0.93).

The majority of cervical screening results positive for HR HPV have normal or NILM cytology [22, 23] and those with HPV 16 or HPV 18 positivity are triaged based on current guidelines [24]. For specimens with NILM cytology, 2 were positive for either HPV 16 or 18 on Alinity m and cobas 4800 in our study, with extended genotyping identifying an additional 2 specimens positive for HPV 45 and 21 specimens positive for other HR HPV genotypes, including 8 positive for Alinity m Other HR HPV A. Two specimens in our cohort with NILM cytology were not detected by cobas 4800 but were positive by Alinity m (Other HR HPV B). Both specimens were also negative on the Aptima HPV assay. We found that the majority of specimens with a negative Alinity m HR HPV result were either normal (NILM) or ASC-US by cytology, and that Alinity m HR HPV positivity increased with the progressive severity of atypical cytology classification, consistent with data presented in a recent epidemiologic report [3].

For specimens with ASC-US cytology, OPA between cobas 4800 and Alinity m for HR HPV Detected results was 94.6% and κ=0.88. Three specimens were positive on cobas (HR HPV Other) but not detected by Alinity m; of these, two specimens were also positive on the Aptima HPV assay. One of these two Aptima-positive samples was positive for HPV 18/45 on the Aptima GT assay and discordant from the cobas 4800 and Alinity m results. These results are not completely unexpected, the ASC-US cytological classification is often inconsistent and subjective, with inadequate reproducible diagnosis. One specimen from a 49-year-old patient classified as LSIL by cytology was positive for HPV16 with cobas 4800 but with a high Ct (39.7) near the cobas 4800 assay cutoff (40.5) [25]. This specimen was not detected by the Alinity m assay or the Aptima GT assay. However, additional histological diagnosis details were available for this specimen, the cervical biopsy showed no transformation zone present, no high-grade dysplasia, and was classified as low-grade squamous intraepithelial lesion/cervical intraepithelial neoplasia 1 (CIN 1). The endocervical curettage was also classified as low-grade squamous intraepithelial lesion/cervical intraepithelial neoplasia 1 (CIN 1). Despite the single discordant case across 3 assays in this cohort with ≥LSIL cytology, OPA between the Alinity m and cobas 4800 assays was 97.3% with a kappa value of 0.89, demonstrating near perfect agreement between the 2 assays.

In this cohort, for specimens with a positive Alinity m HR HPV 16, 18, or Other HR HPV A (31/33/52/58) result, we observed a lower average age for specimens with ≥LSIL cytology (∼40.7 years) versus NILM cytology (∼48.3 years). There was no remarkable difference in the average age for specimens with ≥LSIL (37.2 years) versus NILM (37.3 years) cytology with a positive Alinity m Other HR HPV B (35/39/51/56/59/66/68). Many specimens (n=41/93) with an HPV positive result, regardless of cytology, were in the age group 30-44 years. When comparing HPV-positive specimens from individuals ≤29 years versus ≥30 years, we noted a significantly higher number with NILM and ≥LSIL cytology in the older group and a higher number of ASC-US cytology in the younger group. This finding is consistent with previous reports in the literature.

One specimen that was assessed as cervical adenocarcinoma by cytology was negative on both the cobas 4800 and Alinity m assays. Further workup led to a diagnosis of endometrial carcinoma. As very few cases of endometrial cancer (<10%) are positive for HPV, this case highlights the clinical utility of molecular HPV testing in screening programs to identify cases that are likely to progress to cervical cancer. [26] Five specimens in this cohort had results beyond one genotype or group with the Alinity m HR HPV assay. Three specimens were positive for Other HR HPV A (31/33/52/58) and Other HR HPV B (35/39/51/56/59/66/68), one was NILM, one was ASC-US, and one was LSIL by cytology. One specimen that was LSIL by cytology had an HPV 18 and Other HR HPV B (35/39/51/56/59/66/68) result. The fifth specimen was HSIL by cytology with an HPV 45 and Other HR HPV A (31/33/52/58) result. The association, additional influence, and cumulative effects of multiple type HR HPV co-infections on cervical carcinogenesis is a subject of ongoing research to assess the impact to cervical disease progression.

In contrast to the cobas 4800 assay, which reports the 12 non-16/18 HPV genotypes together, the Alinity m HR HPV assay stratifies HPV 45 and groups the 4 moderately carcinogenic HR HPV genotypes into Other HR HPV A (31/33/52/58) and the 7 less carcinogenic HR HPV genotypes into Other HR HPV B (35/39/51/56/59/66/68). In specimens with ≥LSIL cytology, Alinity m identified six specimens positive for HPV 45 (three of which were classified as HSIL by cytology), nine positive for other HR HPV A, and 14 positive for other HR HPV B genotypes. Notably, HPV 18 was detected by both Alinity m and cobas 4800 in only two samples that were ≥LSIL. The Other HR HPV A genotypes on Alinity m are responsible for 15% of cervical cancers and 11% of all HPV-associated cancers [27]. Although histologic analysis was not available for the specimens in this cohort, previous studies evaluated the use of genotyping stratification similar to the Alinity m HR HPV assay in women with ASC-US cytology. That study found that referring only those with HPV 16, 18, or Other HR HPV A (31/33/52/58) results on Alinity m to colposcopy reduced colposcopy referrals by 37% at the expense of delaying detection of only 8% of ≥CIN2 lesions [28]. Additional investigational study and clinical trial outcomes suggest that extended genotyping will play an enhanced role in clinical practice in the future with the need to stratify higher-risk HPV genotypes to further triage patients beyond the detection of all 14 HR HPV genotypes, particularly for populations with high HPV vaccine coverage [29].

Evolving recommendations favoring incorporation of HR HPV primary screening in cervical cancer screening programs continue to gain momentum in the United States. HR HPV testing is a highly sensitive, efficient, and objective approach to the prevention of cervical cancer that does not depend on the subjective morphological interpretation of cytological results. The high negative predictive value and high sensitivity of molecular HR HPV tests provides higher confidence than cytology alone to identify missed cervical precancerous lesions and additional assurance beyond a negative cytology result of a low risk for ≥CIN2 development [30-33]. Collectively, the application of HR HPV testing offers an advantage over cytology in early detection with a reduction in the future risk of high-grade CIN and cancer as well as extension of the interval between screenings [7]. The Alinity m HR HPV assay is designed to aid in the detection of specimens that have an increased risk of progressing to high-grade disease (HSIL/CIN2+) in screening and co-testing populations. Based on reported cumulative 3-year ≥CIN3 risk profiles for HPV 45, 31, 33, 52, and 58, the extended genotyping capability of Alinity m HR HPV assay offers additional clinical value in improving patient cervical cancer risk stratification. [8-10]

Limitations of the study include the absence of complete biopsy/histology results for determining individual disease status and a limited amount of patient samples in the cohort. We also did not track HPV vaccination status in our study.

## Data Availability

All data produced in the present study are available upon reasonable request to the authors

## Acknowledgements

Stacey Tobin, PhD, provided editorial support for manuscript preparation, with compensation from Abbott Laboratories.

## Author contributions

Joshua Kostera, PhD: conceptualization, formal analysis, funding acquisition, methodology, supervision, writing (original draft, review & editing)

Almedina Tursunovic, MLS (ASCP): data curation, methodology

Paige Botts, MB (ASCP): data curation, methodology

April Davis, MLS (AMT): data curation

Regina Galloway, MT (ASCP): data curation

Tong Yang, MD: conceptualization, data curation, formal analysis, funding acquisition, investigation, methodology, supervision, validation, writing (review & editing)

All authors reviewed the draft and approved the manuscript for submission.

**Funding source:** Consumables and reagents for this study were provided by Abbott Molecular Diagnostics.

## Abbreviations

AGC: atypical glandular cells
ASC-US: atypical squamous cells of undetermined significance
ASC-H: Atypical squamous cells, cannot exclude HSIL
CC: cellular control
CN: cycle number
HPV: human papillomavirus
HR: high risk
HSIL: high-grade squamous intraepithelial lesion
LBC: liquid-based cytology
LSIL: low-grade squamous intraepithelial lesion
NILM: negative for intraepithelial lesions or malignancy
STI: sexually transmitted infection

